# Inequalities in childhood vaccine uptake: a longitudinal analysis of national coverage in England 2019-23

**DOI:** 10.1101/2024.02.03.24301936

**Authors:** Aidan Flatt, Roberto Vivancos, Neil French, Sophie Quinn, Matthew Ashton, Valérie Decraene, Daniel Hungerford, David Taylor-Robinson

**Author notes:** Equal contributors (Daniel Hungerford, David Taylor-Robinson).

## Abstract

**Objective:** This study aims to quantify changes in inequalities in childhood vaccination uptake in the context of steadily declining overall childhood vaccination rates in England.

**Design:** Cross-sectional longitudinal study.

**Setting:** We analysed general practice (GP) level data for five childhood vaccinations (MMR1, MMR2, rotavirus, the pneumococcal (PCV) booster and the six-in-one vaccine) from the Cover of Vaccination Uptake Evaluated Rapidly dataset in England.

**Participants:** Children under 5 years of age eligible for paediatric immunisations between April 2019 and March 2023 registered at GPs in England.

**Main outcome measures:** Changes in quarterly vaccine uptake over time compared by deprivation level. Regression analyses to quantify the change in inequalities in vaccine uptake over time, expressed as changes in the Slope Index of Inequality (SII). We estimated cumulative susceptibility to measles and rotavirus disease at age five.

**Results:** The absolute inequality in vaccine uptake in 2019/20 was largest for MMR2 at 5 years of age (SII -9.8%; 95% CI -9.2 to -10.4). In all vaccinations the SII for uptake increased over the study period: six-in-one -5.1% to -7.8%; rotavirus -7.7% to -10.6%; PCV booster -7.9% to -9.9%; MMR1 at 2 years of age -8.1% to -10.1%, MMR1 -3.3% to -5.9% and MMR2 at 5 years of age -9.8% to -13.7%. The number of measles susceptible children in the least deprived decile increased 15-fold to 20958, and 20-fold to 25345 in the most deprived decile. For rotavirus there was a 14-fold increase in the least deprived decile, and a 16-fold increase in the most deprived decile to 45201.

**Conclusion:** Inequalities in childhood vaccination are increasing in England as uptake rates for five key childhood vaccinations have decreased between 2019 and 2023, below the recommended 95% uptake target. Urgent action is needed to strengthen systems for childhood vaccination, with a key focus on reducing inequalities.

**What is already known on this topic?:**

- Uptake rates of childhood vaccinations in England have been steadily declining in the last decade.
- Socioeconomic deprivation is associated with lower rates of childhood vaccination uptake.

**What this study adds:**

- This analysis of national vaccination coverage data shows decreasing coverage and increasing inequality in five key childhood vaccinations in England from 2019 to 2023.
- The most pronounced increase in inequality over time is seen in the MMR2 vaccination, with a 40% relative increase, whereby the absolute difference in vaccination uptake rates between GP practices serving the lowest and highest levels of deprivation increased from 9.8% to 13.7% across the study period.
- Where vaccination catch up is not implemented, an increasing cumulative number of children more susceptible to infection exists as deprivation increases.
- Policy and practice should respond quickly to address rising socio-economic inequalities in vaccine uptake in children by strengthening systems and tackling the drivers of low vaccination uptake for disadvantaged children.

## Introduction

Vaccination is a foundational public heath intervention, critical for population health and to reduce health inequalities resulting from infectious diseases[1]. However, vaccination uptake rates are affected by socioeconomic factors, with stark inequalities in uptake in many high-income countries [2–7]. Reduced access to and acceptability of childhood vaccinations, with more prevalent vaccine hesitancy in disadvantaged groups, likely to play a role in the generation of these inequalities[8].

According to global studies, barriers to vaccine uptake in socially disadvantaged groups include perceptions of risk, low confidence in vaccinations, distrust of services, barriers to access, lack of community endorsement, and poor communication from trusted providers and community leaders[9,10].

The World Health Organization (WHO) recommends childhood vaccination uptake should exceed a threshold of 95% for effective immunity within a population[11,12]. Vaccination rates in England have declined steadily over the last decade, with few of those included in the routine vaccination schedule reaching overall uptake rates above the 95% threshold[13]. Furthermore, many aspects of health inequalities for children were compounded in England over the period of the covid-19 pandemic[14]. Vaccine-related inequalities were evident both during the covid-19 vaccine rollout[15,16] and post-pandemic, with children growing up in disadvantaged socio-economic circumstances less likely to access immunisations, and more likely to experience worse health outcomes[7,17].

The vaccination schedule in England protects children against fifteen key vaccine preventable diseases and periodically administers vaccinations from 8 weeks to 14 years of age[18]. In England the Cover of Vaccination Evaluated Rapidly (COVER) programme reports vaccination uptake rates in children aged up to 5 years, quarterly and annually. These data are published by the UK Health Security Agency (UKHSA) and are publicly available but have not been assessed from a health equity perspective in the post-covid-19 pandemic era. Understanding how inequalities in vaccination uptake in children are evolving at a small area level in England is essential to inform policy, proactively strengthen public health systems and for the design of effective interventions to reduce inequalities. We therefore aimed to describe the impact of socioeconomic deprivation on the uptake of five key vaccinations included in the childhood immunisation schedule in England (table 1) before, during and after the covid-19 pandemic.

**Table 1.**
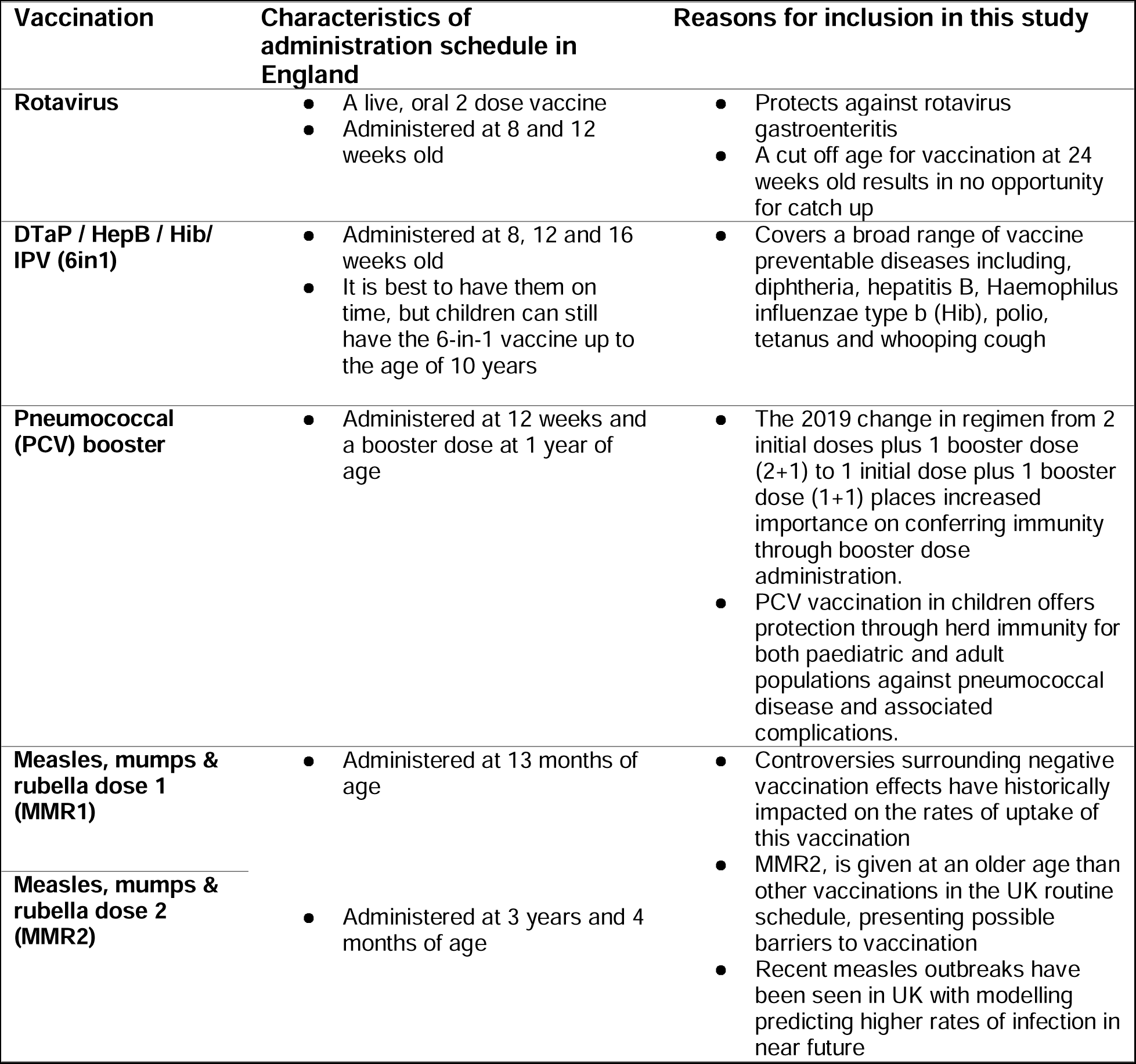
For the five vaccinations chosen to be included in this study, their characteristics of administration and reasons for their inclusion are presented[18–21].

## Methods

### Study design, population and data sources

We analysed longitudinal data captured in the COVER programme in England to assess vaccination uptake rates in children aged up to 5 years[22]. COVER data record how many children registered with a GP in England have received their scheduled immunisations by the age of twelve months, two years and 5 years of age. The data is captured quarterly and annually at GP practice level between April 2019 and March 2023, where quarters cover three-month periods of data collection between April-June, July-September, October-December and January-March.

### Vaccine uptake measures

We extracted uptake rates over time for five childhood vaccinations. The cut off age for analysis was dependent on the vaccination and the detail provided in the COVER data; 6in1 (three completed doses up to first birthday), rotavirus (two completed doses up to first birthday), MMR1 (first dose up to second birthday and fifth birthday), PCV booster (1 dose up to second birthday), and MMR2 (first and second dose up to fifth birthday). We calculated weighted vaccination rates using the uptake rate (%) and number of children in the relevant age group for each vaccination for GP practices in England.

#### Exclusions

Data contained in the COVER programme are provided by Child Health Information Service (CHIS) providers, and completeness of practices and data quality may vary. We excluded practices where the population at the relevant ages had a denominator of less than five. The total number of children excluded did not exceed 1% of the total denominator of the relevant age group. This was the case for all vaccinations analysed. We also excluded GP practices where their identifying code was labelled ‘unknown’. For all vaccinations, we excluded the local authority codes for City of London 714 and Isles of Scilly 906 due to small population sizes. For rotavirus vaccination, we excluded two further local authorities for data derived reasons (Surrey Heartlands 805 and Bradford 209) identified following the investigation of outliers using spaghetti plots.

### Explanatory variables

We measured socioeconomic deprivation using the English Indices of Multiple Deprivation (IMD) scores for each GP practice in England from 2019[4,23]. The IMD is the official measure of relative deprivation at the area level in England. It follows an established methodological framework in broadly defining deprivation to encompass a wide range of an individual’s living conditions. These scores use 39 indicators related to income, employment, health and disability, education, crime, barriers to housing and living environment. We extracted GP level deprivation scores [24], which capture the deprivation of the registered population from the National General Practice Profiles within the Public Health England Fingertips Dashboard utilising population weighting at the Lower Super Output Area (LSOA) level[25,26]. In the descriptive analyses, we categorised deprivation scores into ten deciles, where decile 1 represented 10% of the total number of GP practices in the sample with the least deprivation, and decile 10 contained the 10% of GP practices in the most deprived areas.

### Statistical analysis

We first assessed descriptive trends over time, plotting population weighted uptake of each vaccination by IMD decile. We assessed the absolute difference in vaccination uptake between IMD decile 1 and 10 at the start and at the end of the study period. We describe these as the percentage point difference between the most and least deprived deciles at quarter number 1 (April 2019 to June 2019) and quarter number 16 (January 2023 to March 2023). The time period covering the covid-19 pandemic was added to these plots, between April 2020 and March 2022, which was discerned by when normal service within the NHS was deemed to have resumed and recommendations for covid-19 testing were removed from public policy[27]. We calculated the difference in vaccination uptake rates between two comparable quarters (October-December 2019, and October-December 2022) to assess for possible seasonal influences. We also produced scatterplots for each of the 16 quarters in the time period, plotting total vaccination uptake for each ODS Upper Tier Local Authority against IMD deprivation as a continuous score (supplementary materials). We do not provide detailed estimates of overall uptake by vaccine over time as these data are available in open access COVER reports.

We then undertook regression analyses, using deprivation scores as a continuous measure. To create this continuous measure, we first ranked all included GP practices in order of IMD score for each quarter. We then expressed the population of children in each practice as a proportion of the total number of children across all practices for the quarter. The cumulative number of proportions of children in GP practices of increasing deprivation was calculated from 0 to 1, followed by the mid-point of each range between subsequent GP practices. From this we used the regression to calculate the Slope Index of Inequality (SII), which represents the estimated coefficient of these mid-point values when they are used as a continuous exposure variable in the regression[28,29]. The SII can be interpreted as the absolute difference in vaccination uptake rates between GP practices of lowest and highest levels of deprivation, whilst also accounting for the distribution of the population of children across these GP practices.

To quantify changing inequalities over time we used random effect linear regression models for each of the five vaccinations being investigated. We used vaccination uptake rate as the outcome; the quarter number and weighted deprivation rank as continuous independent variables; and random intercepts and slopes to account for correlations in measurements between GP practice clusters. The model output expressed the SII for each time period. We grouped quarters into four annual years (April to April), resulting in separate regression models to give annual SII values for 2019/20, 2020/21, 2021/22, 2022/23. These values represent the difference in vaccine uptake from least to most deprived. We also assessed the interaction between quarter number and weighted deprivation rank at the 0.05 and 0.95 confidence levels using fixed effects models for each vaccination (supplementary materials section B).

### Estimated numbers susceptible to measles and rotavirus in the study population

We undertook an additional analysis for MMR and rotavirus vaccination to assess the cumulative number of children likely to be susceptible due to lack of vaccination during the study period. We estimated the cumulative number of children susceptible to measles using methodology from Keenan et al.,. [30]. Assuming effectiveness of 97% for a full two dose schedule and 93% for a partial one dose schedule [30]. As the COVER data is cross-sectional, we could only estimate the cumulative number of susceptible children during the study period at five years of age, without consideration of prior infection or catch-up MMR vaccination occurring post data collection.

Therefore, the analysis is likely to overestimate the true number of children susceptible to measles for this study population. Susceptible numbers were calculated using the following formula:

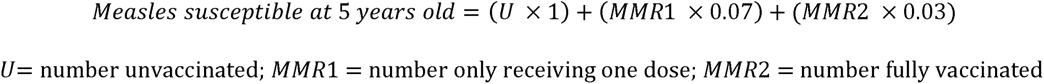

Given the cut off vaccine eligibility of six months of age for administration of the rotavirus vaccination, there is not an opportunity for catch-up of unvaccinated individuals. To estimate susceptibility for rotavirus, we used COVER data combined with vaccine effectiveness estimates from the literature of 87% for a full two dose vaccine schedule and 72% for partial schedule (first dose)[31]. Because COVER only provides numerators for full dose rotavirus coverage at one year of age we estimated the number of individuals receiving one dose using an assumption that an additional 5% of those eligible in the denominator would have received just one dose, and the remainder are considered unvaccinated [32,33]. Susceptible numbers were calculated using the following formula:

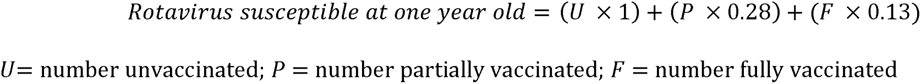

We plotted the theoretical cumulative population of children susceptible to rotavirus disease and measles based on GP practice level denominators. We then stratified this by IMD decile to illustrate the difference in the number of estimated susceptible children across the levels of population deprivation. All analyses were undertaken in RStudio V1.4.1717.

### Patient and public involvement

No patients or members of the public were directly involved in this piece of research. However, our research programme into equity in vaccine use and outcomes has been informed by patients through our institute’s patient public involvement and engagement panel. We have also held a series of consultation groups with parents/carers on equity and communication around immunisations, which addressed the benefits, concerns, barriers, and priorities and informed how the results are presented in this paper. The findings for this study have and will be shared with public health organisations and presented at regional and national events, with health, lay and government representation.

## Results

### Trends in vaccination uptake

Between April 2019 and March 2023, the mean number of GPs included in the study per quarter was 6557 for all vaccinations accept rotavirus (n=6374) (supplementary table S1). Over the study period, 2 386 317 (2 309 674 for rotavirus vaccination) eligible children at one year, 2 456 020 at two years and 2 689 304 at five years were included in the study. The total overall uptake fell for all vaccinations, ranging between 0.1 percentage points (pp) for the 6in1 vaccine and 1.6pp for MMR1 at five years (supplementary table S2). The highest vaccine uptake was for MMR1 at five years in April 2020 to June 2020 at 95.0% and lowest was for MMR2 at five years in April 2022 to June 2022 (84.9%).

Over the study period uptake fell short of the WHO 95% target for all vaccines studied across all IMD deciles except for the top three least deprived deciles for the 6in1 vaccination (fig 1). For all vaccinations, the absolute difference in uptake between the least and most deprived deciles increased over the study period. For the 6in1 vaccination, the absolute difference in vaccination uptake between the least and most deprived deciles in the starting quarter was 3.3% and increased to 7.4% (+4.1pp) by the final quarter of the data collection period. For rotavirus vaccination, there was an increase in the absolute difference from 6.3% to 9.1% (+2.8pp). For the PCV booster vaccination, the absolute difference increased from 5.6% to 8.6% (+3pp). For MMR1 at two years, from 5.8% to 8.3% (+2.5pp), and for MMR2 at five years, from 5.3% to 11.5% (+6.2pp)

**Fig 1.**
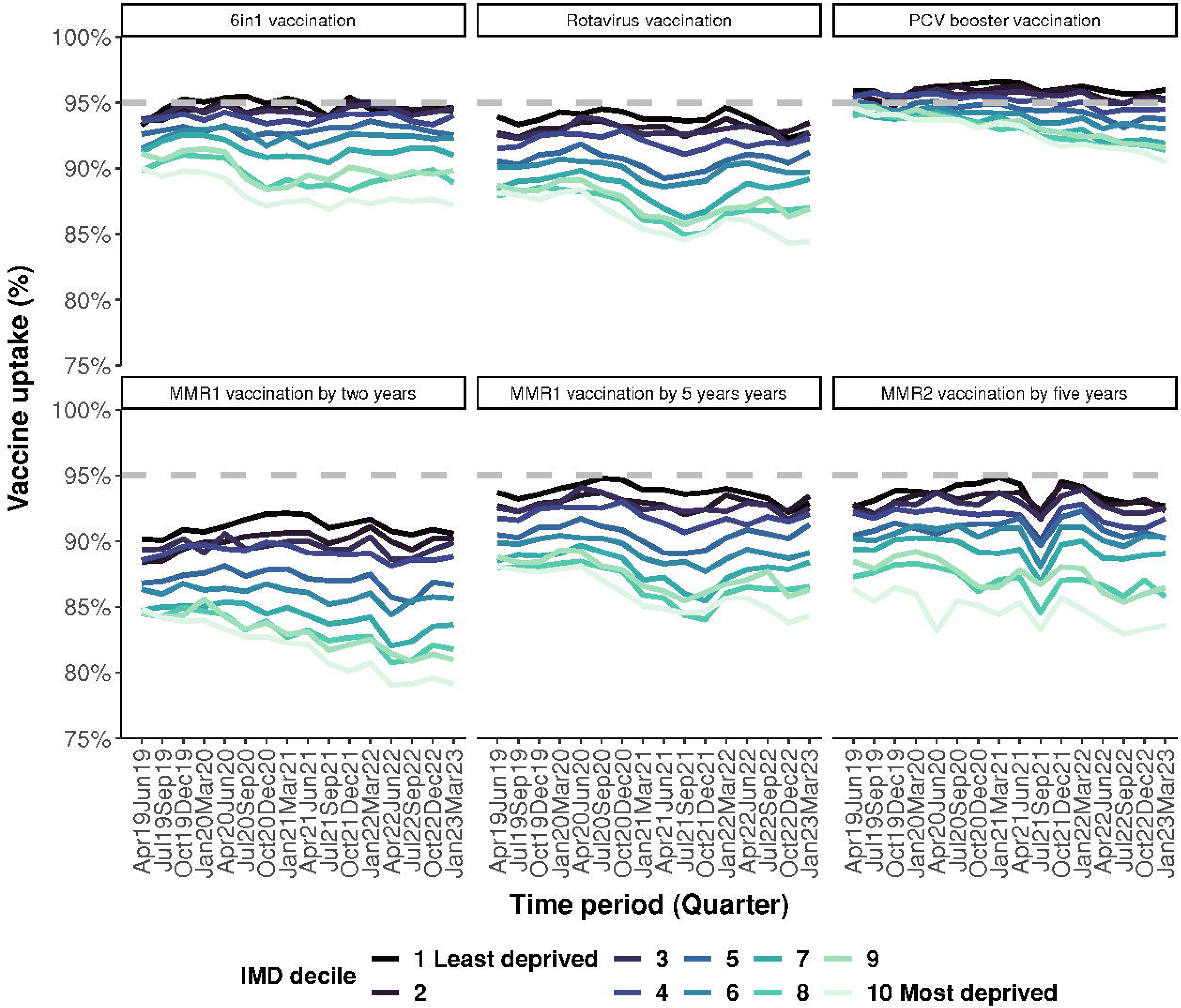
Population weighted uptake of each vaccination studied over time, stratified by Index of Multiple Deprivation (IMD) decile. The grey box represents the period covering the covid-19 pandemic. The grey dashed line represents the WHO 95% vaccine uptake target.

To account for possible seasonal factors relating to trends in vaccination uptake, the absolute difference in vaccination uptake between IMD decile 1 and 10 was calculated for two comparable quarters (October-December 2019 and October-December 2022). These results are shown in supplementary materials table S2. For all vaccinations, the drop in percentage uptake between 2019 and 2022 was greater in those in the most deprived IMD decile compared to the least deprived. Uptake of MMR1 at five years and MMR2 at five years marginally increased in the least deprived decile, by 0.1pp and 0.4pp.

Figure 2 shows the changing slope index of inequality over the study period from the regression models and summarises the SII result from the annual linear regression models calculated. For full model outputs with confidence intervals, refer to supplementary section B. All vaccinations under study have a baseline SII in 2019/20, but the size of the SII varies by vaccine type (fig 2 and supplementary table S3). The SII for vaccine uptake at baseline was largest for MMR2 (-9.8%; 95% CI -9.2 to -10.4) and smallest for the MMR1 at five years (-3.3% 95% CI -2.9 to -3.7). In all vaccinations the SII for vaccine uptake increases from 2019/2020 to 2020/21, then again from 2020/21 to 2021/22. For rotavirus vaccination, MMR1 at five years and MMR2 at five years point estimates for the SII for vaccination uptake continues to increase between 2021/22 and 2022/23 (fig 2).

**Fig 2.**
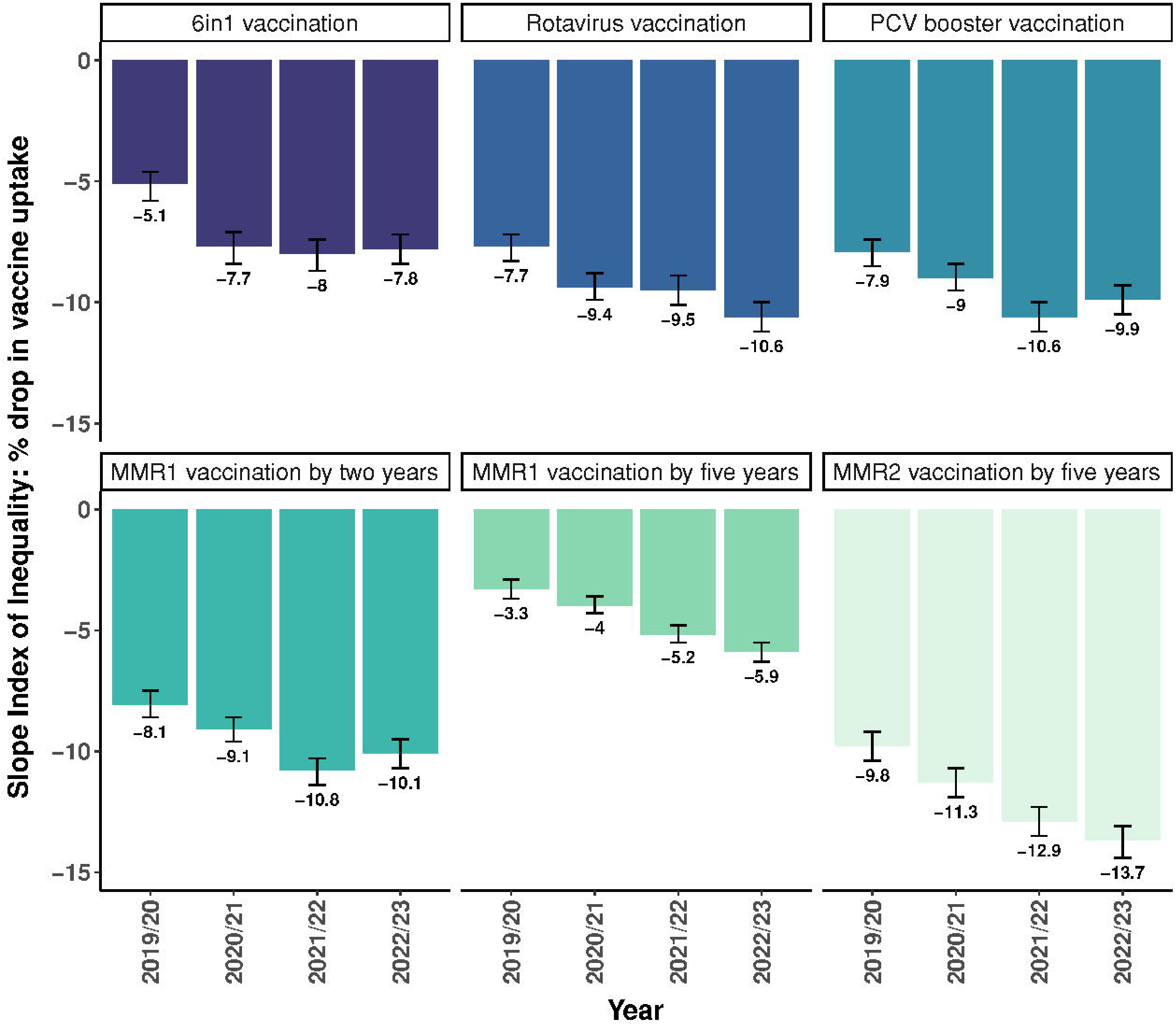
Bar charts for each of the vaccinations analysed showing the drop in vaccine uptake % from least to most deprived as represented by the Slope Index of Inequality (SII).

### Cumulative susceptibility

For measles infection the estimated cumulative number of susceptible children aged 5 years was 1364 in the least deprived decile rising 15-fold to 20 958 by the end of the study period. In the most deprived decile, there was a 20-fold change over the study period, from 1296 to 25 345 (fig 3). For rotavirus disease, the estimated cumulative number of susceptible children aged 1 year at the start of the study period was 2292 in the least deprived decile, rising 14-fold to 32 981 by the end of the study period. In the most deprived decile, there was a 16-fold-change over the study from to 45 201 (fig 3).

**Fig 3.**
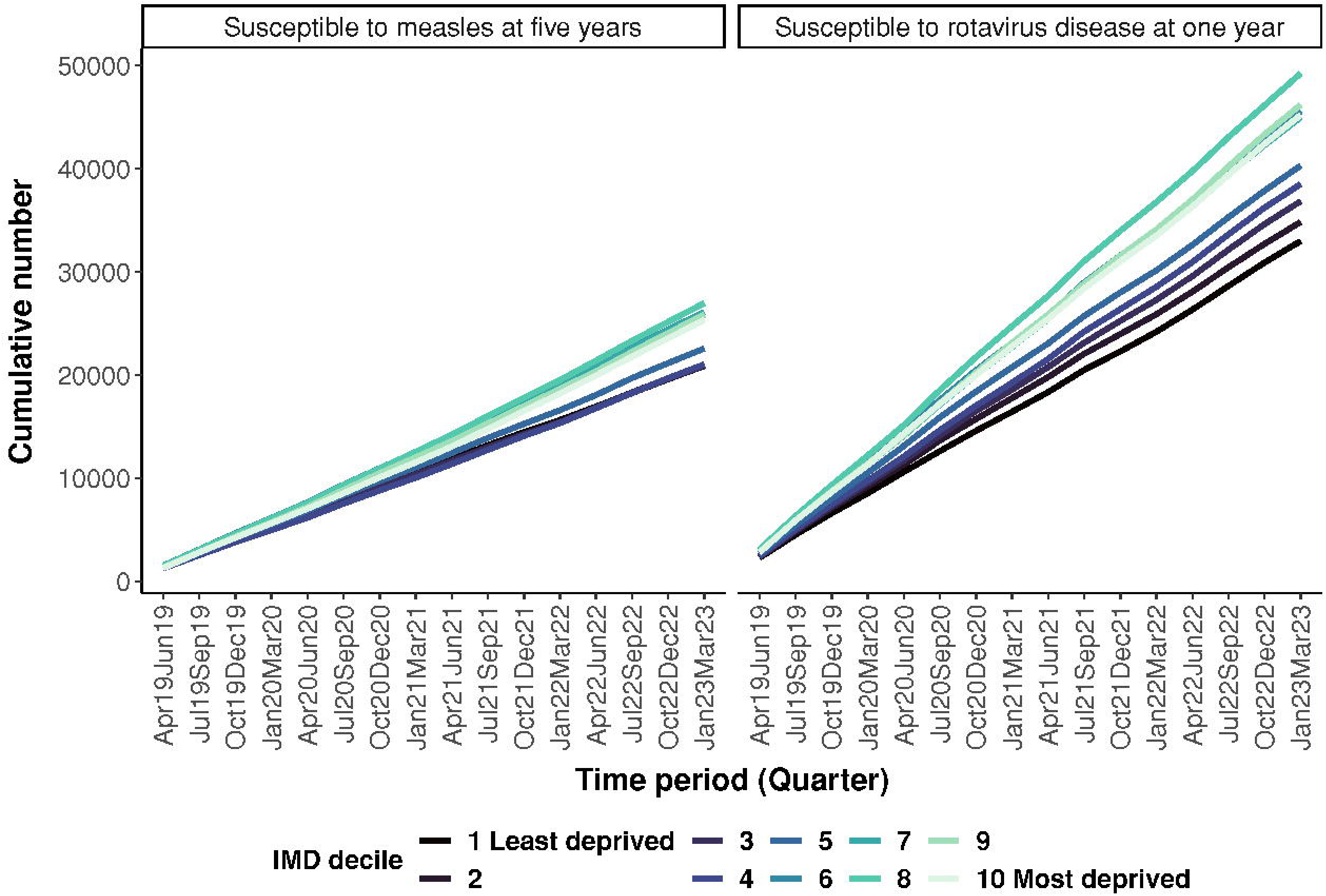
The number of children accumulating over the study period that are likely to be susceptible to rotavirus disease or measles infection, by Index of Multiple Deprivation decile.

### Robustness tests

For the outputs from the sensitivity analysis undertaken to assess the effect of excluding Surrey and Bradford from the rotavirus vaccination analysis (see supplemental materials). Excluding these local authorities due to data derived anomalies did not have a significant effect on the regression analysis and gives confidence in the robustness of the final analysis undertaken.

## Discussion

This England-wide analysis of GP level paediatric vaccine uptake highlights stark socio-economic inequalities, whereby children living in areas of higher deprivation consistently have lower uptake rates of vaccinations up to the age of five years. We show increasing inequality in vaccine uptake between 2019 and 2023. The most pronounced increase in inequality over time is seen in the MMR2 vaccination, with a 40% relative increase, whereby the absolute difference in vaccination uptake rates between GP practices serving the lowest and highest levels of deprivation increased from 9.8% to 13.7% across the study period. For all paediatric vaccinations studied, the uptake rates in England did not exceed the WHO recommended 95% threshold in the more deprived populations.

### Findings in context

Paediatric vaccine uptake has decreased globally in the wake of the COVID-19 pandemic [34], with an estimated 20.5 million children worldwide either unvaccinated or under-vaccinated in 2022.[35] Confidence in childhood vaccinations is at an alarmingly low level across European and Central Asian regions[36]. Childhood vaccination rates have shown some post-pandemic recovery,[37], though as evidenced in our study uptake remains lower than pre-pandemic levels.

Few studies have assessed trends in inequalities of paediatric vaccines over this period [29], and our study highlights the dramatic widening of inequalities in England. This is a critical public health issue, as more deprived areas often have higher population density, more frequent overcrowding in the home, poorer baseline health and higher rates of comorbidity[38]. These conditions increase the risks of infectious disease transmission, outbreaks and poorer health outcomes[39]. Therefore, the effects of falling vaccine uptake will not be felt equally across populations. Furthermore, as evidenced post-Wakefield, broken trust surrounding vaccination and healthcare is harder to rebuild in more deprived population groups, and risks amplifying existing health inequalities[21]. Beyond these general patterns, there are specific implications for falling uptake of each of the vaccinations studied here, and the diseases they protect against.

For MMR herd immunity, the WHO recommends 95% coverage of two doses of MMR owing to the highly infectious nature of measles [20,40]. This threshold has historically not been reached in England[4], with our study showing that this is now unmet by over 15% in the most deprived populations. Measles cases have begun to rise in the UK and Europe, with modelling predicting tens of thousands of cases in London alone in coming months[20]. In early 2024, there have been measles outbreaks in large urban areas in England. In Birmingham 216 confirmed and 103 probable cases were detected between October 2023 and 18^th^ January 2024 and a national incident declared by UKHSA [41].

Our study shows a reduction in uptake of the PCV booster, most pronounced in more deprived populations. This is in the context of the schedule switch in 2019, from two primary doses and one booster dose (2L+L1) to one primary and a booster (1 + 1) schedule (table 1). The booster dose is even more critical for protection in the new schedule and therefore widening inequality is concerning for disease in disadvantaged adults who require herd protection and where the risk of serious illness and invasive pneumococcal disease is disproportionately higher [19,42]. Furthermore, the risk of pneumonia is disproportionately higher in children living in areas of increased deprivation [43].

We show the largest fall in rotavirus vaccination uptake since its introduction to the UK schedule in 2013. Prior to vaccine introduction, rotavirus was the leading cause of acute gastroenteritis in children, with hospital admissions highest in more deprived populations[33,44]. Rotavirus vaccination significantly reduced these admissions with high vaccine effectiveness[31,45] and also reduced inequalities in disease burden [33]. This was despite lower rotavirus vaccine uptake in more deprived groups, as also shown in our study. Rotavirus vaccination eligibility ends at 6 months of age, meaning there is no opportunity for catch up[46]. This makes the growing inequity in uptake and cumulative increase of susceptible children particularly concerning.

Increasing inequalities in uptake rates of the 6in1 vaccination presents concerns for several vaccine preventable diseases (DTaP/Hib/HepB/IPV). Recent detection of variant poliovirus on environmental surveillance in England, has increased the risk of infection, outbreaks and clinical poliomyelitis[47]. There have also been widespread increases in pertussis (whooping cough) in England in 2023, which could be partial attributed to falling vaccine uptake but also due to waning immunity in older children and adults, compounded by reduced exposure to natural infections during the covid-19 pandemic[48].

### Strengths and limitations

This study examines uptake of childhood vaccinations across England, utilising temporal, small area-level data. As such, it provides a responsive and detailed picture and allows for timely decision-making surrounding interventions. These data are publicly available and are released quarterly, so analyses can be repeated and tailored for local needs.

Our analyses are predominately descriptive and rely on aggregated routine health data. We are unable to investigate the mechanisms and processes that could explain why socioeconomic inequalities in childhood vaccine uptake have increased. We also are unable to account for all potential confounders or other explanatory factors. Social deprivation is only one factor that influences unequal vaccine uptake, and others include, disability, gender, ethnicity, religion, geography and age. In addition, evidence suggests that migrants, travellers, prisoners, and being looked after child all influence vaccine inequalities not just for overall coverage, but also for timing of vaccines and completion of vaccine schedules[8].

Data limitations also exist within this study, including incorrectly recorded uptake rates for rotavirus vaccination uptake in some areas. These data anomalies were examined in a sensitivity analysis and were not deemed to significantly impact the findings. While these data capture whether children have received their eligible vaccine doses, the specific date of receipt is unknown. These data do not include children who are not registered at GP practices or capture vaccinations delivered in private settings. Catch-up of vaccinations outside of the routine paediatric immunisations is also not captured in these data. Furthermore, without access to individual level records for the whole population it is not possible to use these data to accurately assess susceptibility in the paediatric and adult populations.

### Implications for policy and practice

Giving every child the best start in life is recognised as being critical to narrowing health inequalities. Childhood vaccination is potentially a powerful “levelling-up” intervention [49]. NHS England has a legal duty to offer immunisation to ‘hard to reach groups’ and the core service specification for the National Immunisation Programme drawn up between NHS and public health bodies has reduction in health inequalities as a key objective in delivery of the programme[8]. The broad principle of health equity action requires action on the upstream social drivers of ill health and inequalities[50]. The Marmot Review, introduced the concept of ‘proportionate universalism’, suggesting that health equity actions must be universal, not targeted, but with a scale and intensity that is proportionate to the level of disadvantage[49].

Systems strengthening through rapid investment and effective partnerships between stakeholders and institutions including Integrated Care Systems (ICS), Public Health Departments, the UKHSA, NHS England and academic institutions is required[51]. Promising approaches likely involve strengthening and investment at a local level in supplementary outreach services designed to meet specifics needs of underserved populations. These services should be integrated in a network incorporating local commissioners, Public Health Departments, Voluntary, Community and Social Enterprise (VCSE), Health and Wellbeing Alliance, primary care and early years settings, drawing on insights from services, community leaders and neighbourhood level data [52]. In addition, knowledge exchange between the public sector and industry will allow adoption of innovative technologies to improve immunisation delivery in both routine preventative care and outbreak response.

Partnerships will only be able to act efficiently when real-time data on local population immunisation status and susceptibility are routinely available to local public health teams. Area level secure Data Environments aimed at mobilising data for public health analytics were used to evaluate pandemic responses and vaccination uptake[53]. However, these systems are not mature across England for any imminent outbreak or prevention response. Robust local analytics will help focus interventions on improving vaccination uptake at the time of children’s eligibility within the routine schedule. As catch-up interventions are costly, challenging and not available for all vaccinations, meaning that missed vaccination creates increasing sized pools of susceptible children as deprivation increases. We therefore should also be concerned about the build of up of susceptible post-school teenagers and young adults. The current increases in whooping cough and measles cases in England are likely to herald more widespread outbreaks.

## Conclusion

We have demonstrated stark and increasing inequalities in childhood vaccination uptake over recent years in England. Overall rates of vaccine uptake for five key childhood vaccinations have declined between 2019 and 2023, with more rapid declines observed with increasing levels of deprivation. Vaccine uptake was below the recommended 95% WHO uptake target throughout the study period for all vaccinations. These findings strongly support the urgent need for effective vaccination systems strengthening, proportionate to levels of need, in addition to interventions and catch-up campaigns in underserved populations.

## Supporting information

RECORD Research Checklist

## Ethics statements

### Ethical approval

Not required as data used for this study are anonymised, aggregated and publicly available at https://www.gov.uk/government/collections/vaccine-uptake#cover-of-vaccination-evaluated-rapidly-programme

### Data availability statement

No additional data available. All data is open access and available through original sources at the UK Health Security Agency https://www.gov.uk/government/collections/vaccine-uptake#cover-of-vaccination-evaluated-rapidly-programme.

## Acknowledgements

The authors acknowledge and thank the COVER data and Public Health Outcomes Framework data for its accessibility and use for research. The authors thank Dr Davara Bennett for support with the SII analysis methodology code. The authors also thank the GP practices involved in submitting vaccination data.

## Contributors

DTR, DH, RV, AF conceptualised the study. AF acquired the data. AF and DH carried out the statistical analyses. NF, DH, DTR, RV, SQ, AF contributed to methodology. DH and DTR contributed to supervision. AF and DH carried out visualisation. AF, DH, DTR writing – original draft preparation. DH, DTR, AF, SQ, VD, NF, MA, RV contributed to writing-reviewing and editing. The corresponding author attests that all listed authors meet authorship criteria.

