## Supplementary Materials for "Inequalities in childhood vaccine uptake: a longitudinal analysis of national coverage in England 2019-23"

Supplementary material

### Supplementary section A: additional descriptive tables

Table S1: Number of eligible children and number of GPS included within the dataset per vaccination and quarter, April 2019 to March 2023

| Quarter | 6in1 vaccination at 1 year of age | | Rotavirus vaccination at 1 year of age | | PCV booster vaccination at 2 years of age | | MMR1 at 2 years of age | | MMR1 at 5 years of age | | MMR2 at 5 years of age | |
| --- | --- | --- | --- | --- | --- | --- | --- | --- | --- | --- | --- | --- |
|  | **GPs (n)** | **Eligible children (n)** | **GPs (n)** | **Eligible children (n)** | **GPs (n)** | **Eligible children (n)** | **GPs (n)** | **Eligible children (n)** | **GPs (n)** | **Eligible children (n)** | **GPs (n)** | **Eligible children (n)** |
| Apr19 Jun19 | 6840 | 155266 | 6650 | 150264 | 6840 | 160992 | 6840 | 160992 | 6840 | 167672 | 6840 | 167672 |
| Jul19 Sep19 | 6807 | 160955 | 6618 | 155845 | 6807 | 168777 | 6807 | 168777 | 6807 | 176470 | 6807 | 176470 |
| Oct19 Dec19 | 6738 | 154413 | 6549 | 149352 | 6738 | 161071 | 6738 | 161071 | 6738 | 169588 | 6738 | 169588 |
| Jan20 Mar20 | 6733 | 146119 | 6544 | 141529 | 6733 | 151533 | 6733 | 151533 | 6733 | 162955 | 6733 | 162955 |
| Apr20 Jun20 | 6656 | 150643 | 6468 | 145840 | 6656 | 156354 | 6656 | 156354 | 6656 | 168768 | 6656 | 168768 |
| Jul20 Sep20 | 6641 | 161359 | 6453 | 156439 | 6641 | 165197 | 6641 | 165197 | 6641 | 179760 | 6641 | 179760 |
| Oct20 Dec20 | 6612 | 149245 | 6424 | 144551 | 6612 | 156216 | 6612 | 156216 | 6612 | 171565 | 6612 | 171565 |
| Jan21 Mar21 | 6580 | 142880 | 6393 | 138346 | 6580 | 146924 | 6580 | 146924 | 6580 | 164343 | 6580 | 164343 |
| Apr21 Jun21 | 6519 | 144034 | 6332 | 139534 | 6519 | 151708 | 6519 | 151708 | 6519 | 169058 | 6519 | 169058 |
| Jul21 Sep21 | 6426 | 147268 | 6243 | 142537 | 6426 | 156170 | 6426 | 156170 | 6426 | 172796 | 6426 | 172796 |
| Oct21 Dec21 | 6428 | 141459 | 6250 | 136770 | 6428 | 149645 | 6428 | 149645 | 6428 | 165365 | 6428 | 165365 |
| Jan22 Mar22 | 6401 | 137447 | 6225 | 132954 | 6401 | 143928 | 6401 | 143928 | 6401 | 158315 | 6401 | 158315 |
| Apr22 Jun22 | 6400 | 145702 | 6225 | 140913 | 6400 | 146470 | 6400 | 146470 | 6400 | 165183 | 6400 | 165183 |
| Jul22 Sep22 | 6402 | 156297 | 6226 | 151119 | 6402 | 151907 | 6402 | 151907 | 6402 | 172434 | 6402 | 172434 |
| Oct22 Dec22 | 6380 | 152289 | 6206 | 147271 | 6380 | 146848 | 6380 | 146848 | 6380 | 167378 | 6380 | 167378 |
| Jan23 Mar23 | 6354 | 140941 | 6180 | 136410 | 6354 | 142280 | 6354 | 142280 | 6354 | 157654 | 6354 | 157654 |
| Mean | 6557 | 149145 | 6374 | 144355 | 6557 | 153501 | 6557 | 153501 | 6557 | 168082 | 6557 | 168082 |
| Total Apr19 to Mar23 |  | 2386317 |  | 2309674 |  | 2456020 |  | 2456020 |  | 2689304 |  | 2689304 |

Table S2: Percentage uptake of five routine childhood immunisation, comparison between quarters in the most and least deprived deciles and overall, April 2019 to March 2023

| IMD decile | Vaccine uptake by quarter (%) | | | | | | | | | | | | | | | |
| --- | --- | --- | --- | --- | --- | --- | --- | --- | --- | --- | --- | --- | --- | --- | --- | --- |
|  | **Apr19 Jun19** | **Jul19 Sep19** | **Oct19 Dec19** | **Jan20 Mar20** | **Apr20 Jun20** | **Jul20 Sep20** | **Oct20 Dec20** | **Jan21 Mar21** | **Apr21 Jun21** | **Jul21 Sep21** | **Oct21 Dec21** | **Jan22 Mar22** | **Apr22 Jun22** | **Jul22 Sep22** | **Oct22 Dec22** | **Jan23 Mar23** |
|  | 6in1 vaccination at 1year of age | | | | | | | | | | | | | | | |
| 1 Least deprived | 93.3 | 94.4 | 95.3 | 95.0 | 95.4 | 95.5 | 94.9 | 95.3 | 94.9 | 93.9 | 95.4 | 94.5 | 94.7 | 94.2 | 94.7 | 94.6 |
| 10 Most deprived | 90.0 | 89.4 | 89.8 | 89.7 | 89.3 | 87.8 | 87.1 | 87.5 | 87.5 | 86.9 | 87.6 | 87.3 | 87.7 | 87.5 | 87.6 | 87.2 |
| Overall | 92.1 | 92.4 | 93.0 | 92.8 | 93.0 | 92.1 | 91.7 | 92.0 | 91.8 | 91.6 | 92.3 | 92.3 | 92.2 | 92.1 | 92.1 | 92.0 |
|  | Rotavirus vaccination at 1 year of age | | | | | | | | | | | | | | | |
| 1 Least deprived | 92.6 | 93.1 | 93.8 | 93.8 | 93.6 | 94.3 | 94.4 | 94.8 | 94.3 | 91.7 | 94.5 | 94.1 | 93.2 | 93.0 | 93.0 | 92.7 |
| 10 Most deprived | 86.3 | 85.4 | 86.4 | 86.0 | 83.2 | 85.4 | 85.1 | 84.4 | 85.3 | 83.3 | 85.7 | 84.9 | 83.8 | 82.9 | 83.3 | 83.6 |
| Overall | 90.2 | 90.0 | 90.8 | 90.9 | 90.6 | 90.6 | 90.2 | 90.4 | 90.6 | 88.5 | 90.8 | 90.9 | 89.8 | 89.3 | 89.7 | 89.5 |
|  | PCV booster vaccination at 2 years of age | | | | | | | | | | | | | | | |
| 1 Least deprived | 93.7 | 93.2 | 93.6 | 94.0 | 94.3 | 94.8 | 94.7 | 93.9 | 93.9 | 93.6 | 93.7 | 94.0 | 93.7 | 93.3 | 92.2 | 92.9 |
| 10 Most deprived | 88.1 | 87.8 | 87.7 | 87.9 | 88.1 | 87.1 | 86.1 | 85.1 | 84.9 | 84.5 | 84.6 | 85.7 | 85.7 | 84.9 | 83.8 | 84.3 |
| Overall | 90.5 | 90.3 | 90.6 | 90.9 | 91.3 | 90.9 | 90.6 | 89.5 | 89.3 | 88.7 | 88.8 | 89.7 | 89.9 | 89.8 | 89.1 | 89.8 |
|  | MMR1 at 2 years of age | | | | | | | | | | | | | | | |
| 1 Least deprived | 93.9 | 93.3 | 93.7 | 94.3 | 94.1 | 94.5 | 94.3 | 93.8 | 93.8 | 93.6 | 93.7 | 94.6 | 93.9 | 93.1 | 92.3 | 92.7 |
| 10 Most deprived | 88.1 | 88.0 | 87.6 | 88.2 | 88.4 | 87.1 | 86.2 | 85.4 | 85.0 | 84.6 | 85.1 | 86.2 | 86.0 | 85.2 | 84.3 | 84.4 |
| Overall | 90.5 | 90.4 | 90.7 | 91.0 | 91.3 | 90.9 | 90.6 | 89.7 | 89.4 | 89.0 | 89.4 | 90.3 | 90.3 | 90.1 | 89.6 | 90.1 |
|  | MMR1 at 5 years of age | | | | | | | | | | | | | | | |
| 1 Least deprived | 95.9 | 95.9 | 95.2 | 96.0 | 96.2 | 96.3 | 96.5 | 96.6 | 96.5 | 95.8 | 96.0 | 96.3 | 95.9 | 95.7 | 95.7 | 96.0 |
| 10 Most deprived | 94.6 | 94.0 | 94.1 | 93.6 | 93.7 | 93.7 | 93.1 | 93.5 | 93.2 | 92.3 | 91.6 | 91.8 | 91.6 | 91.5 | 91.2 | 90.5 |
| Overall | 95.0 | 94.9 | 94.5 | 94.9 | 95.0 | 94.7 | 94.6 | 94.7 | 94.6 | 94.1 | 94.0 | 94.1 | 93.5 | 93.4 | 93.6 | 93.4 |
|  | MMR2 at 5 years of age | | | | | | | | | | | | | | | |
| 1 Least deprived | 90.2 | 90.1 | 90.9 | 90.7 | 91.1 | 91.6 | 92.1 | 92.1 | 92.0 | 91.0 | 91.3 | 91.7 | 90.8 | 90.5 | 90.9 | 90.6 |
| 10 Most deprived | 84.9 | 84.2 | 83.9 | 84.0 | 83.3 | 82.7 | 82.7 | 82.3 | 82.1 | 80.7 | 80.1 | 80.7 | 79.1 | 79.1 | 79.6 | 79.1 |
| Overall | 86.9 | 86.8 | 87.3 | 87.3 | 87.4 | 87.0 | 87.3 | 87.1 | 86.9 | 86.1 | 86.3 | 86.7 | 85.3 | 85.3 | 86.0 | 85.9 |

### Supplementary section B: linear regression model outputs and Slope Index of Inequality SII

Table S3: Annual linear regression model estimates of the Slope Index of Inequality by surveillance year and vaccination, from April 2019 to March 2023

| Time period | Slope Index of Inequality by vaccination type (% and 95% CIs) | | | | | |
| --- | --- | --- | --- | --- | --- | --- |
|  | **6in1** | **Rotavirus** | **PCV booster** | **MMR1 at 2 years** | **MMR2 at 5 years** | **MMR1 at 5 years** |
| 2019/20 | -5.1  (-4.6 to -5.8) | -7.7  (-7.2 to -8.3) | -7.9  (-7.4 to -8.5) | -8.1  (-7.5 to -8.6) | -9.8  (-9.2 to -10.4) | -3.3  (-2.9 to -3.7) |
| 2020/21 | -7.7  (-7.1 to -8.4) | -9.4  (-8.8 to -9.9) | -9.0  (-8.4 to -9.5) | -9.1  (-8.6 to -9.6) | -11.3  (-10.7 to -11.9) | -4.0  (-3.6 to -4.3) |
| 2021/22 | -8.0  (-7.4 to-8.7) | -9.5  (-8.9 to -10.1) | -10.6  (-10.0 to -11.2) | -10.8  (-10.3 to -11.4) | -12.9  (-12.3 to -13.5) | -5.2  (-4.8 to -5.5) |
| 2022/23 | -7.8  (-7.2 to -8.4) | -10.6  (-10.0 to -11.2) | -9.9  (-9.3 to -10.5) | -10.1  (-9.5 to -10.7) | -13.7  (-13.1 to -14.4) | -5.9  (-5.5 to -6.3) |

6in1 vaccination linear regression model outputs:

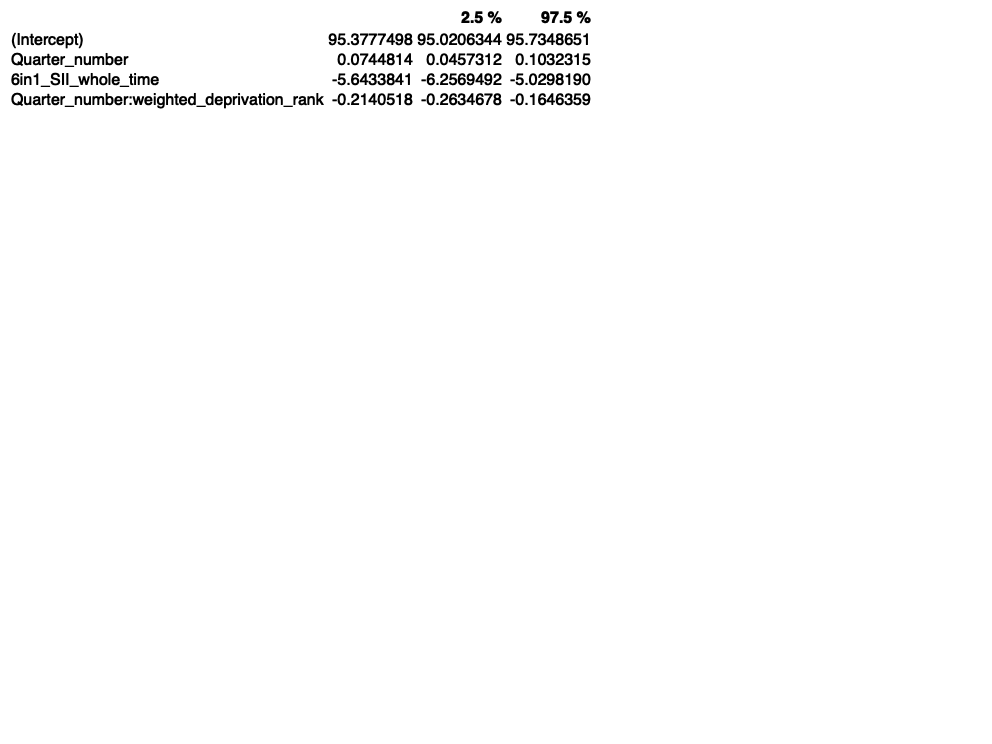

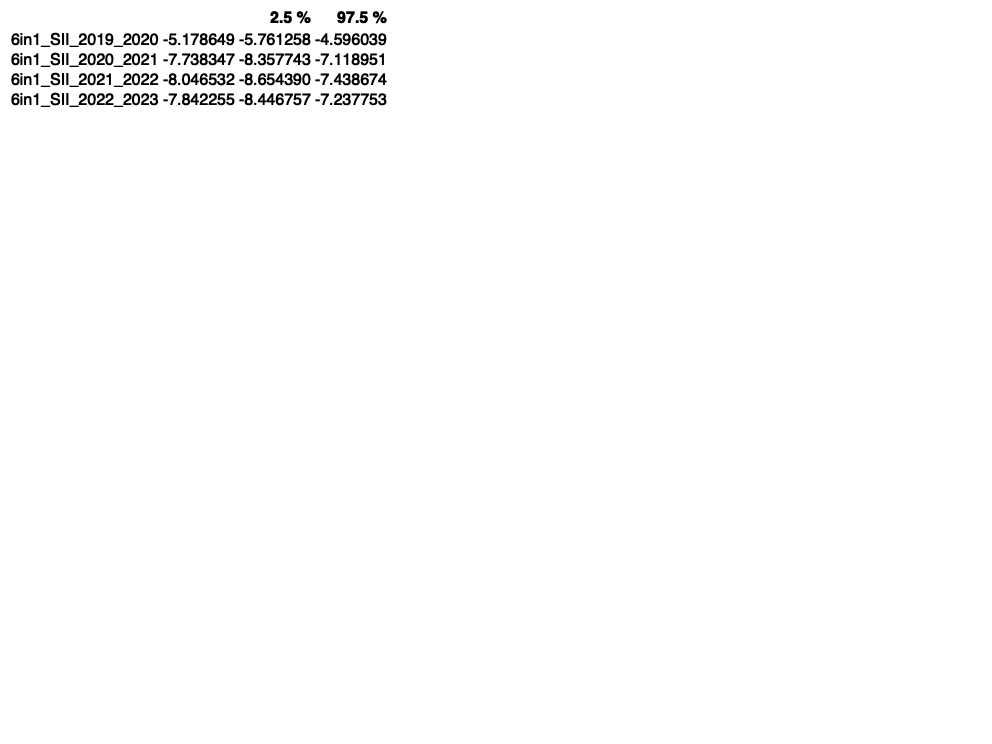

Rotavirus vaccination linear regression model outputs

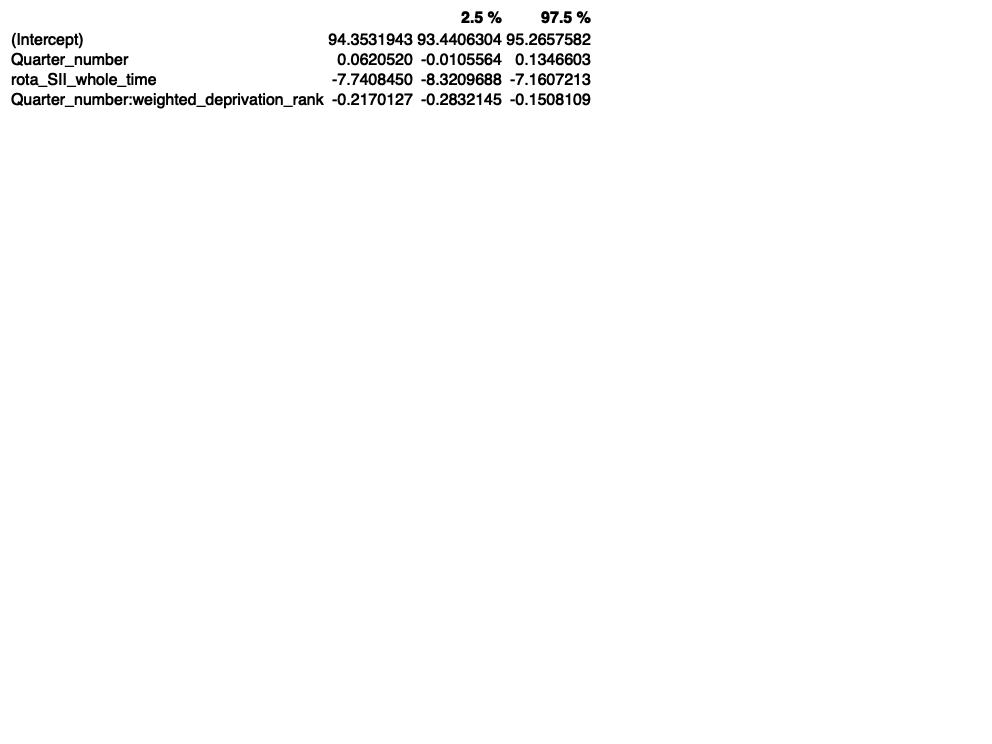

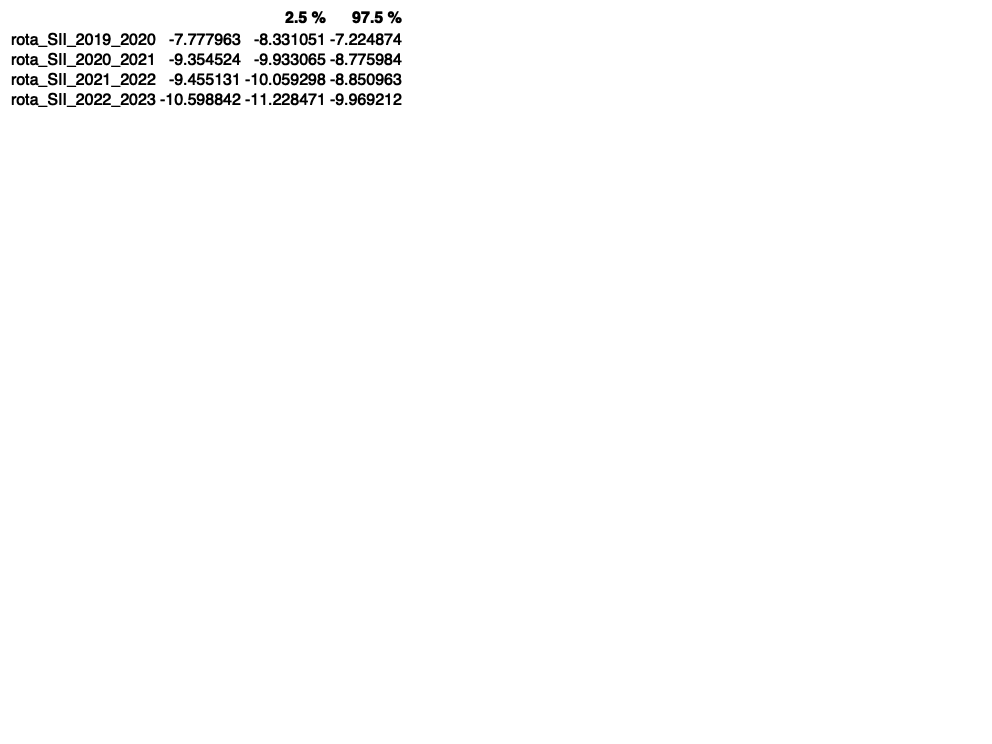

PCV booster vaccination linear regression model outputs:

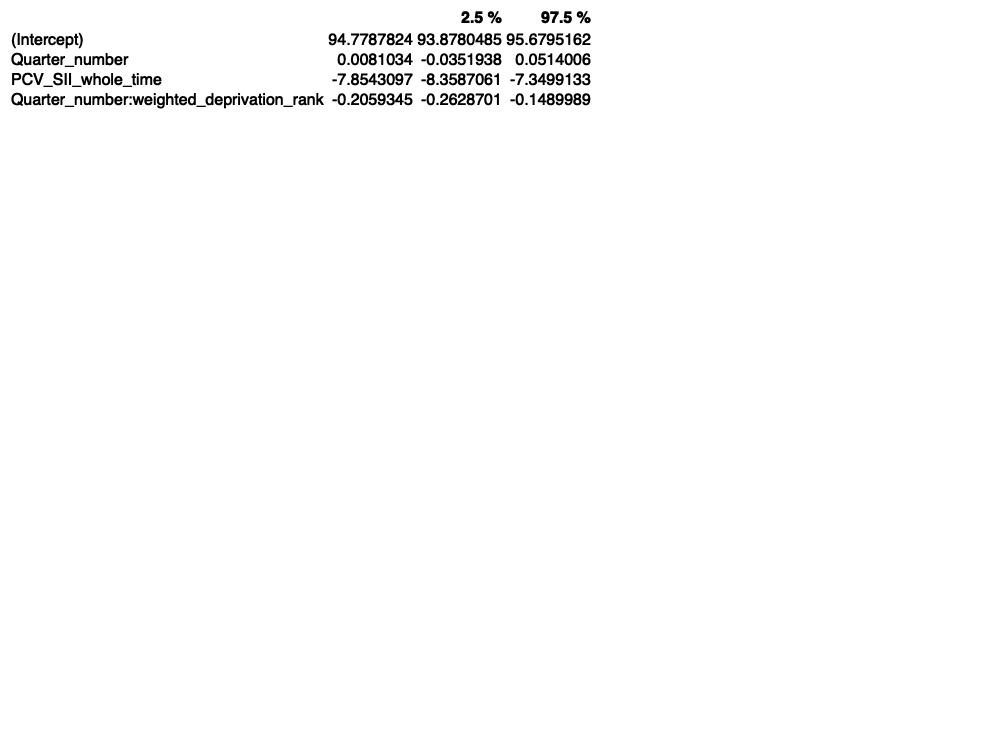

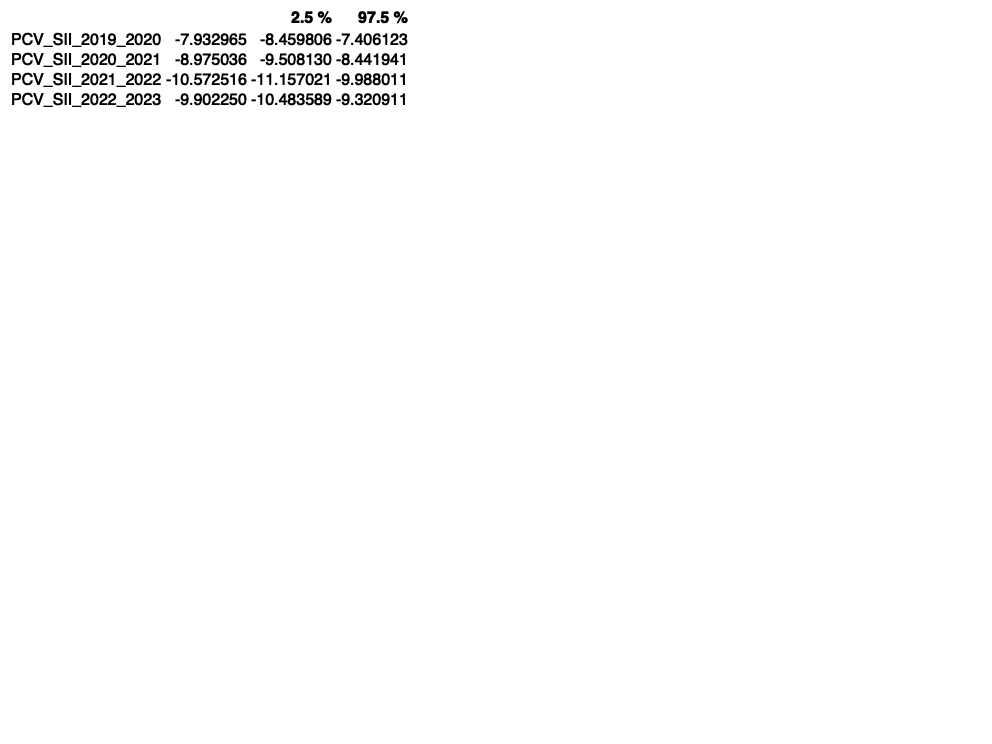

MMR1 at 2 years linear regression model outputs:

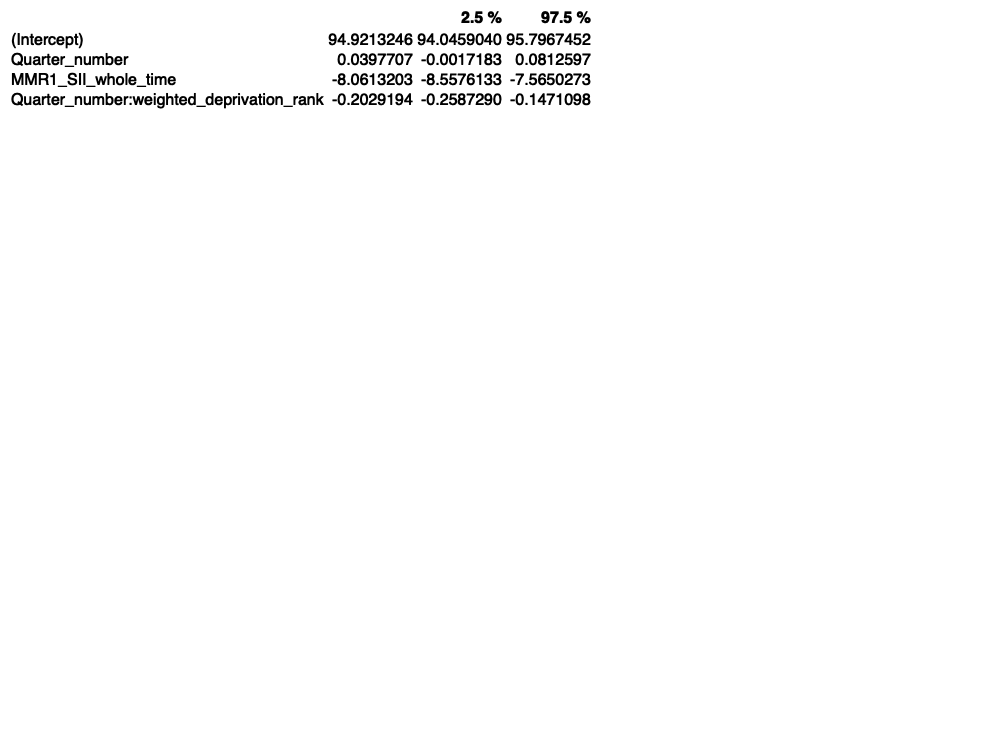

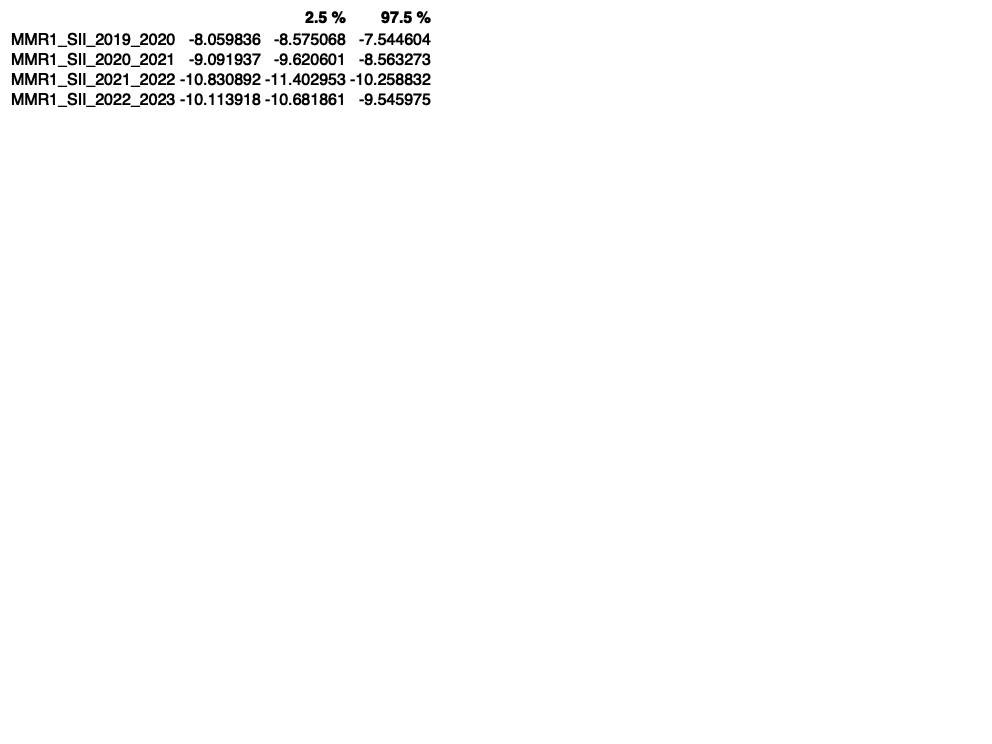

MMR1 at 5 years linear regression model outputs:

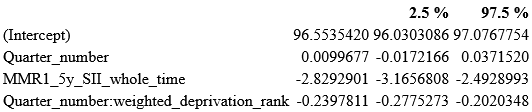

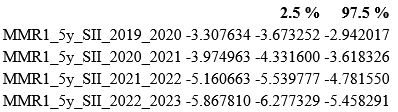

MMR2 at 5 years linear regression model outputs:

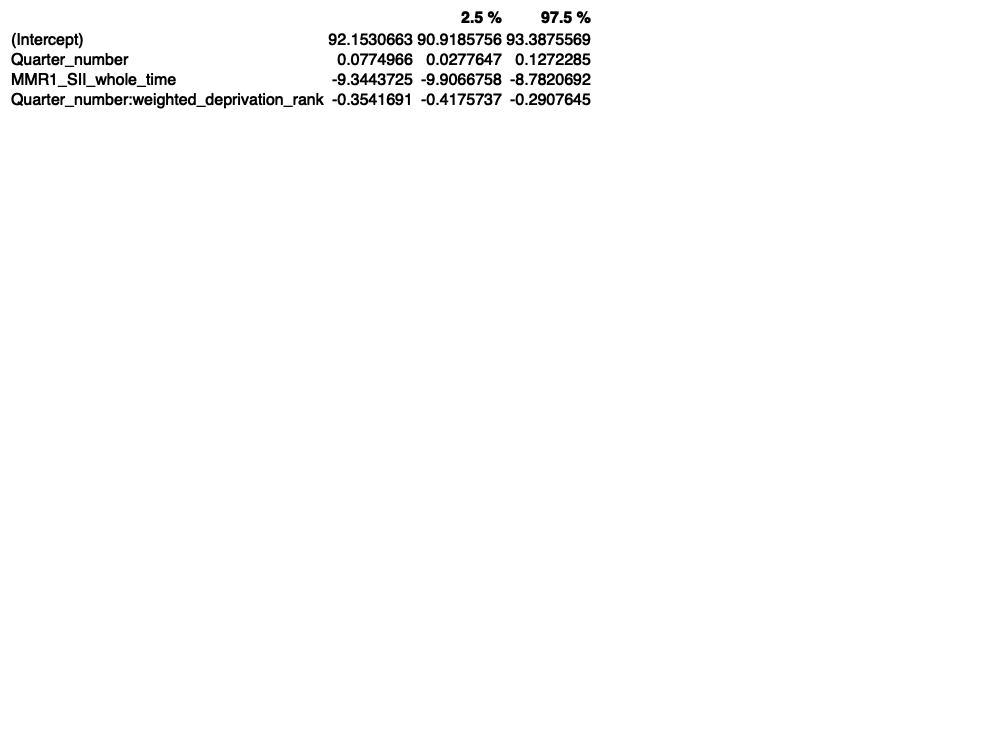

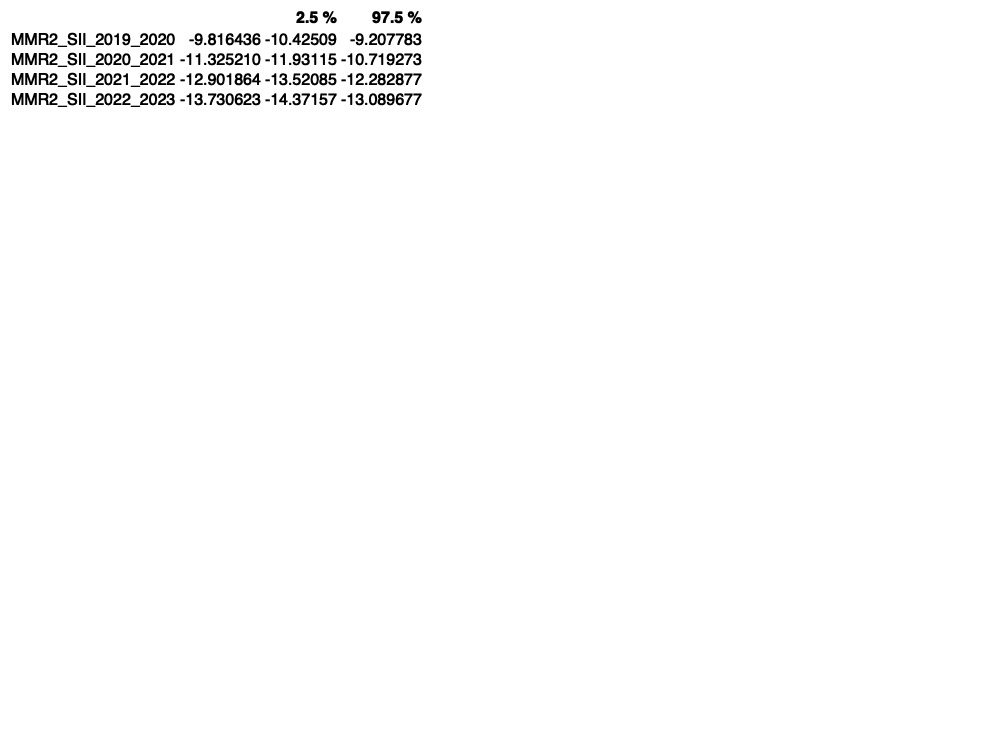

Sensitivity analysis results for rotavirus vaccination:

*Bradford and Surrey not excluded:*

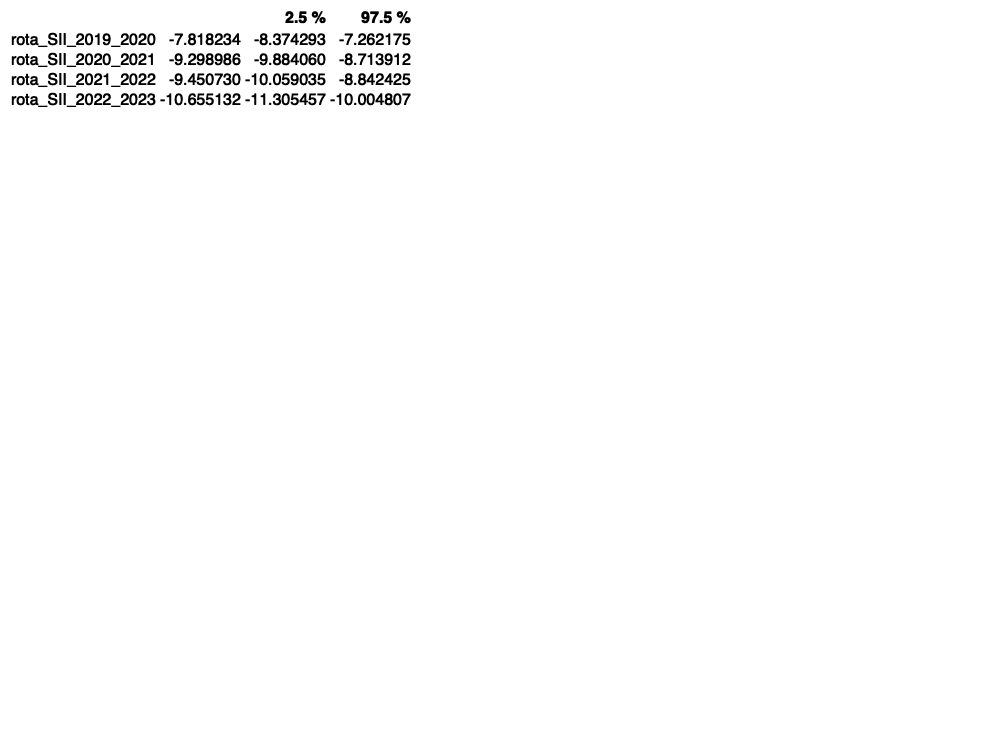

*Bradford and Surrey excluded:*

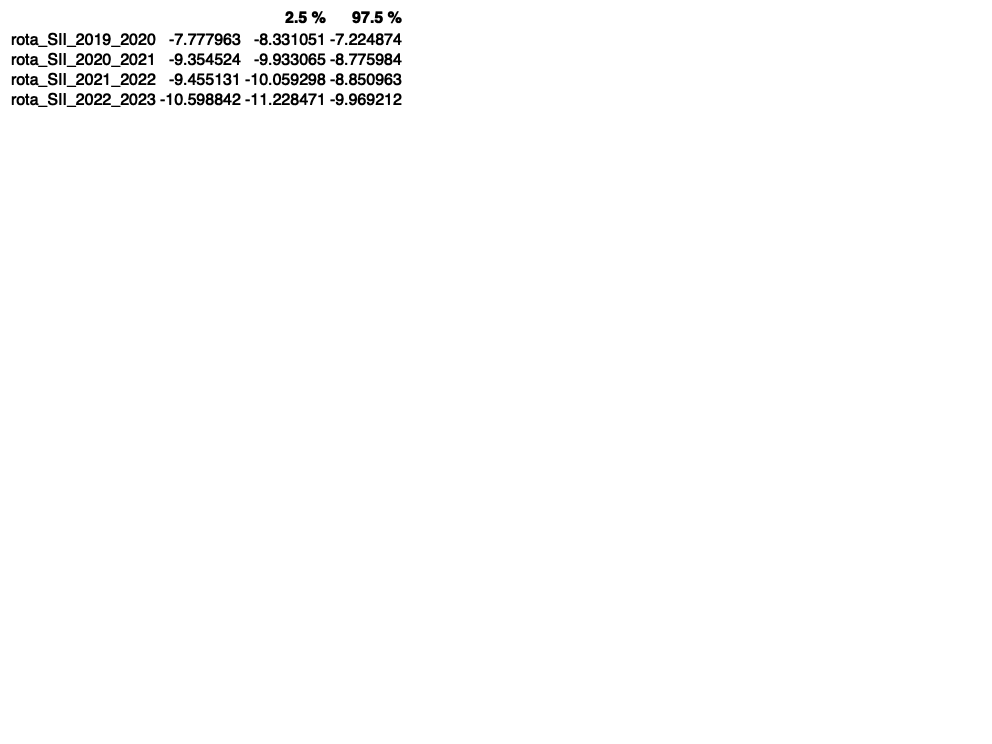
